# Geospatial analysis of the presence of hypertension, diabetes, obesity, and lifestyles in the Peruvian population

**DOI:** 10.1101/2024.08.11.24311820

**Authors:** Joan A. Loayza-Castro, Lupita Ana Maria Valladolid-Sandoval, Luisa Erika Milagros Vásquez Romero, Fiorella E. Zuzunaga-Montoya, Jhonatan Roberto Astucuri Hidalgo, Víctor Juan Vera-Ponce

## Abstract

**Introduction:** In Peru, the prevalence of diabetes mellitus (DM), hypertension (HTN), and obesity has increased significantly while healthy lifestyles have declined. However, the geographic distribution of these conditions at the provincial level has not been fully elucidated.

**Objective:** To conduct a geospatial analysis to provide a comprehensive view of the distribution of DM, HTN, obesity, and lifestyles across Peruvian provinces.

**Materials and Methods:** An analytical cross-sectional study was conducted using data from the 2023 Demographic and Family Health Survey (DHS). Descriptive analysis of the study variables was performed, and geospatial analysis techniques, including heat maps and the Local Moran’s Index, were used to identify distribution patterns and clusters.

**Results:** The national prevalence of DM, HTN, and obesity was 5.66%, 20.43%, and 27.67%, respectively. It was found that 8.35% were current smokers, 2.24% consumed alcohol excessively, and 90.69% consumed fewer than five servings of fruits and vegetables daily. Geospatial analysis revealed hotspots of high DM prevalence in the provinces of Oyón (Lima) and Daniel Alcides Carrión (Pasco), HTN in Putumayo (Loreto) and Corongo (Ancash), and obesity in Putumayo and Maynas (Loreto), and Pallasca (Ancash). Regarding lifestyles, clusters of high alcohol consumption were identified in the provinces of Piura and Cajamarca, high tobacco consumption in Loreto and San Martín, and low fruit and vegetable consumption in Lima and Huánuco.

**Conclusions:** Marked geographical disparities were observed in the prevalence of chronic diseases and unhealthy lifestyles among Peruvian provinces, with a notable concentration in jungle regions and some coastal Andean areas.

## Introduction

Non-communicable chronic diseases (NCDs) represent a global public health challenge with a disproportionate impact on low- and middle-income countries ^(1)^. In Peru, the burden of these diseases has increased significantly in recent decades, reflecting an accelerated epidemiological transition ^(2)^. Diabetes mellitus (DM), hypertension (HTN), and obesity have emerged as priority public health problems, with estimated prevalences of 7.3%, 24.9%, and 22%, respectively ^(3–5)^. However, these aggregated data at the national level mask significant disparities between regions and population groups.

The geographic distribution of NCDs in Peru is intrinsically linked to socioeconomic, environmental, and cultural factors that vary considerably throughout the country. Rapid urbanization, changes in dietary patterns, and the adoption of sedentary lifestyles have contributed to increased risk factors for these diseases ^(6)^. In parallel, lifestyles, including alcohol and tobacco consumption and dietary habits, show complex patterns that reflect cultural traditions and globalization’s influences ^(7)^.

In this context, geospatial analysis is crucial for understanding the distribution and determinants of NCDs and health behaviors in Peru. This methodology identifies high-prevalence clusters, reveals geographic disparities in disease burden, and provides insights into the contextual factors influencing population health ^(8)^. In a country characterized by its geographic and cultural diversity, such as Peru, this approach is precious for informing public health policies adapted to local realities ^(9)^.

Despite the importance of this topic, there is a scarcity of studies that comprehensively examine the geospatial distribution of NCDs and lifestyles in Peru using representative national data. The present study seeks to fill this gap through an exhaustive geospatial analysis of the prevalence of DM, HTN, obesity, and lifestyle patterns in Peru, using data from the 2023 Demographic and Family Health Survey (DHS). Our specific objectives are: 1) to map the geographic distribution of these health conditions and behaviors at the provincial level and 2) to identify clusters of high and low prevalence using advanced spatial analysis techniques.

## Methods

### Study Design

An analytical cross-sectional study using a geospatial approach used secondary data from Peru’s 2023 DHS. DHS is a nationally representative population-based survey designed to provide estimates at the national level by area of residence (urban and rural) and by natural region (Coast, Highlands, and Jungle) ^(10)^.

### Population, Sample, and Eligibility Criteria

DHS employs a probabilistic, stratified, and multi-stage sampling method, ensuring the representativeness of the Peruvian population.

For this study, individuals aged 18 years or older were included. Participants with incomplete data for the main study variables and pregnant women were excluded from the obesity analysis. To ensure reliable blood pressure data, measurement ranges based on previous research were established: between 70 mmHg and 270 mmHg for Systolic Blood Pressure (SBP) and between 50 mmHg and 150 mmHg for Diastolic Blood Pressure (DBP). Measurements outside these ranges were discarded from the study ^(11)^.

### Variables and Measurement

The main variables of this study were DM, HTN, obesity, and lifestyles:

1. Lifestyles: Determined using dimensions of smoking status, alcohol consumption, and fruit and vegetable consumption

a) Smoking status: Obtained from participant self-report and classified as never smoked, former smoker, current smoker, and daily smoker. For geospatial analysis, this was dichotomized into current smokers or not.
b) Alcohol consumption: Based on the survey, self-report and classified as never consumed or not consumed alcohol in the last 12 months, non-excessive consumption in the last 30 days, and excessive consumption (≥ one occasion in the last 30 days and ≥ five drinks for men or ≥ four drinks for women) ^(12,13)^. For geospatial analysis, this was dichotomized into non-consumer/non-excessive versus excessive.
c) Fruit and vegetable consumption: Consider whether consumption was more significant than 5 daily portions ^(14)^.
2. DM: Determined from the question asked to participants: "Has a doctor ever diagnosed you with high blood sugar?"
3. HTN: Evaluated considering SBP measurements ≥ 140 mmHg and/or DBP ≥ 90 mmHg; and/or a history of high blood pressure diagnosis.
4. Obesity: Evaluated using Body Mass Index (BMI) to distinguish participants with obesity (BMI ≥ 30 kg/m²) ^(15)^.

Additional covariables included: sex (female or male), age (in years), natural region (Metropolitan Lima, Rest of Coast, Highlands, and Jungle), educational level (none, primary, secondary, and higher), marital status (single and partnered), wealth index (very poor, poor, medium, rich, and very rich), area of residence (urban and rural), and Height above mean sea level (MASL) (0 to 499 MASL, 500 to 1499 MASL, 1500 to 2999MASL, and 3000 or more MASL).

### Procedures

Data collection followed internationally standardized and validated protocols. Trained personnel from Peru’s National Institute of Statistics and Informatics conducted interviews and anthropometric measurements in selected households.

Daily calibrated anthropometric equipment was used for BMI measurement (weight/height squared). Weight was determined using electronic scales (SECA brand, model 874) with 0.1 kg precision. Participants were weighed in light clothing without shoes. Height was measured with portable stadiometers (SECA brand, model 213) with 1 mm precision, following the Frankfurt protocol for head position. Each measurement was performed in duplicate, using the average for final calculations. A third measurement was taken in case of discrepancies greater than 0.5 kg in weight or 1 cm in height.

Blood pressure was measured using clinically validated automatic digital sphygmomanometers (OMRON brand, model HEM-7130). Two cuff sizes were used to adapt to participants’ arm circumference. Measurements were taken after the participant had been seated and at rest for at least 5 minutes, with the right arm supported at heart level. Two readings were taken with a two-minute interval between them. A third measurement was taken if the difference between readings was greater than 10 mmHg for systolic or 6 mmHg for diastolic pressure. The final value was calculated as the average of the two (or three) readings.

Lifestyle data, including tobacco, alcohol, fruit, and vegetable consumption, were collected through a structured questionnaire administered by trained interviewers. To minimize recall bias, standardized interview techniques were used, and visual aids were employed for estimating fruit and vegetable portion sizes.

Geographic information was obtained from Global Positioning System (GPS) coordinates of sample clusters, recorded during fieldwork. These coordinates were subsequently linked to participant data using unique identifiers, maintaining the confidentiality of individual information.

## Data Analysis

The statistical program R, version 4.3.1, was used. First, a descriptive analysis of patients’ sociodemographic and clinical characteristics was performed. Categorical variables were summarized using absolute and relative frequencies, expressed as percentages. Numerical variables were expressed as means with their respective standard deviations (SD).

Subsequently, a geospatial analysis was conducted to evaluate the geographic distribution of DM, HTN, Obesity, and lifestyles. Prevalences of each of these conditions were calculated by province and then integrated with the shapefile of Peruvian provinces. Heat maps were created using ggplot2 to visualize the distribution of prevalences.

To determine spatial autocorrelation at the provincial level, both global and local Moran’s indices were used, with a significance level of 0.05 using 999 permutations. The permutations generate a null distribution of spatial autocorrelation values considering the hypothesis of no spatial clustering, which allows calculation of p-values and evaluation of the statistical significance of identified spatial patterns. Correlation visualization was performed using a scatter plot with values from -1 to +1, allowing assessment of whether the units of analysis presented positive autocorrelation, negative autocorrelation, or randomization.

Spatial representation was performed using the Local Moran’s Index (LISA), which allowed identification of five types of clustering: 1) provincial clusters with high prevalences of DM, HTN, Obesity, and lifestyles surrounded by provinces with above-average prevalence ("high-high" or hot spots); 2) provincial clusters with high prevalences surrounded by provinces with below-average prevalences ("high-low"); 3) provincial clusters with low prevalences surrounded by provinces with above-average prevalences ("low-high"); 4) provincial clusters with low prevalences surrounded by provinces with low prevalences ("low-low"); and finally 5) provinces with prevalences that have no significant correlation with surrounding provinces ("not significant").

### Ethical Aspects

This study was developed using the DHS survey, which is publicly accessible ^(12)^. The information used does not have any personal specification of the participants, and there was no contact with them. Due to this, a review by a research ethics committee was not considered pertinent. However, in developing this study, the principles of bioethics and the agreements determined in the Declaration of Helsinki were taken into account.

## Results

A total of 33,103 people participated in the study. Considering the main study variables, the prevalence of DM, HTN, and obesity was reported as 5.66%, 20.43%, and 27.67%, respectively. Regarding lifestyles, 8.35% were current smokers, 2.24% consumed alcohol excessively, and 9.31% consumed fewer than 5 servings of fruits and vegetables daily.

The female sex was the most representative, with 52.02% of the samples, 44% of participants were between 36 and 59 years old, and 37.42% lived in Metropolitan Lima. Regarding educational level, 44.29% indicated having secondary education, while 38.87% reported being single, and 18.36% of participants belonged to the very poor category according to the wealth index. 81.78% lived in urban areas, only 1.36% reported having a physical disability, and 66.81% of participants indicated living in geographic areas with an altitude of 0 to 499 MASL.

Figure 1 shows heat maps of the prevalence of DM, HTN, and obesity among participants. In map A, the darker shades of purple represent the highest prevalences of DM, found in the provinces of Castilla (Arequipa) and Puno (Puno), followed by regions in the jungle and north of the country, primarily in the provinces of Mariscal Ramón Castilla (Loreto), Tahuamanu (Madre de Dios), Piura and Ayabaca (Piura), Contralmirante Villar (Tumbes), and Oyon (Lima). Map B shows the prevalence of HTN, with the highest prevalences in the provinces of Putumayo (Loreto) and Corongo (Ancash). Finally, map C represents obesity prevalences, with the darkest green shades in the provinces of Tahuamanu and Manu (Madre de Dios), Talara and Sechura (Piura), Pisco and Palma (Ica), Cañete (Lima), and Camaná (Arequipa).

**Table 1.** Descriptive characteristics of the study sample.

| Characteristic | n = 33,103 |
| --- | --- |
| **Sex** |  |
| Female | 17,222 (52.02%) |
| Male | 15,882 (47.98%) |
| **Age (mean ± DS)** | 40.59 (15.99) |
| **Natural region** |  |
| Metropolitan Lima | 12,386 (37.42%) |
| Resy of coast | 8,569 (25.89%) |
| Mountain Range | 7,907 (23.89%) |
| Jungle | 4,24 (12.81%) |
| **Educational Level** |  |
| No Level | 53 (0.16%) |
| Primary | 6,194 (19.29%) |
| Secondary | 14,218 (44.29%) |
| Superior | 11,639 (36.25%) |
| **Civil status** |  |
| Single | 9,725 (38.87%) |
| With a partner | 15,292 (61.13%) |
| **Wealth index** |  |
| The poorest | 6,077 (18.36%) |
| Poor | 6,546 (19.77%) |
| Medium | 6,809 (20.57%) |
| Rich | 6,923 (20.91%) |
| Richest | 6,749 (20.39%) |
| **Area of residence** |  |
| Urban | 27,072 (81.78%) |
| Rural | 6,031 (18.22%) |
| **Physical disability** |  |
| No | 32,652 (98.64%) |
| Yes | 451 (1.36%) |
| **Arterial Hypertension** |  |
| No | 26,340 (79.57%) |
| Yes | 6,763 (20.43%) |
| **T2DM** |  |
| No | 31,187 (94.34%) |
| Yes | 1,869 (5.66%) |
| **Body Mass Index** |  |
| Normal Weight | 10,323 (31.19%) |
| Overweight | 13,617 (41.14%) |
| Obesity | 9,158 (27.67%) |
| **Altitude (MASL)** |  |
| 0 to 499 | 22,116 (66.81%) |
| 500 to 1499 | 2,576 (7.78%) |
| 1500 to 2999 | 3,700 (11.18%) |
| 3000 or more | 4,711 (14.23%) |
| **Smoker Status** |  |
| Has never smoked | 27,315 (82.52%) |
| Former smoker | 2,575 (7.78%) |
| Currently smoker | 2,763 (8.35%) |
| Daily smoker | 449 (1.36%) |
| **Alcohol consumption** |  |
| Never or did not use in the last 12 months | 8,963 (27.08%) |
| Non-excessive | 23,398(70.68%) |
| Excessive | 743 (2.24%) |
| **Fruit and vegetable consumption** |  |
| < 5 servings per day | 30,022 (90.69%) |
| ≥ 5 servings per day | 3,081 (9.31%) |

**Figure 1.**
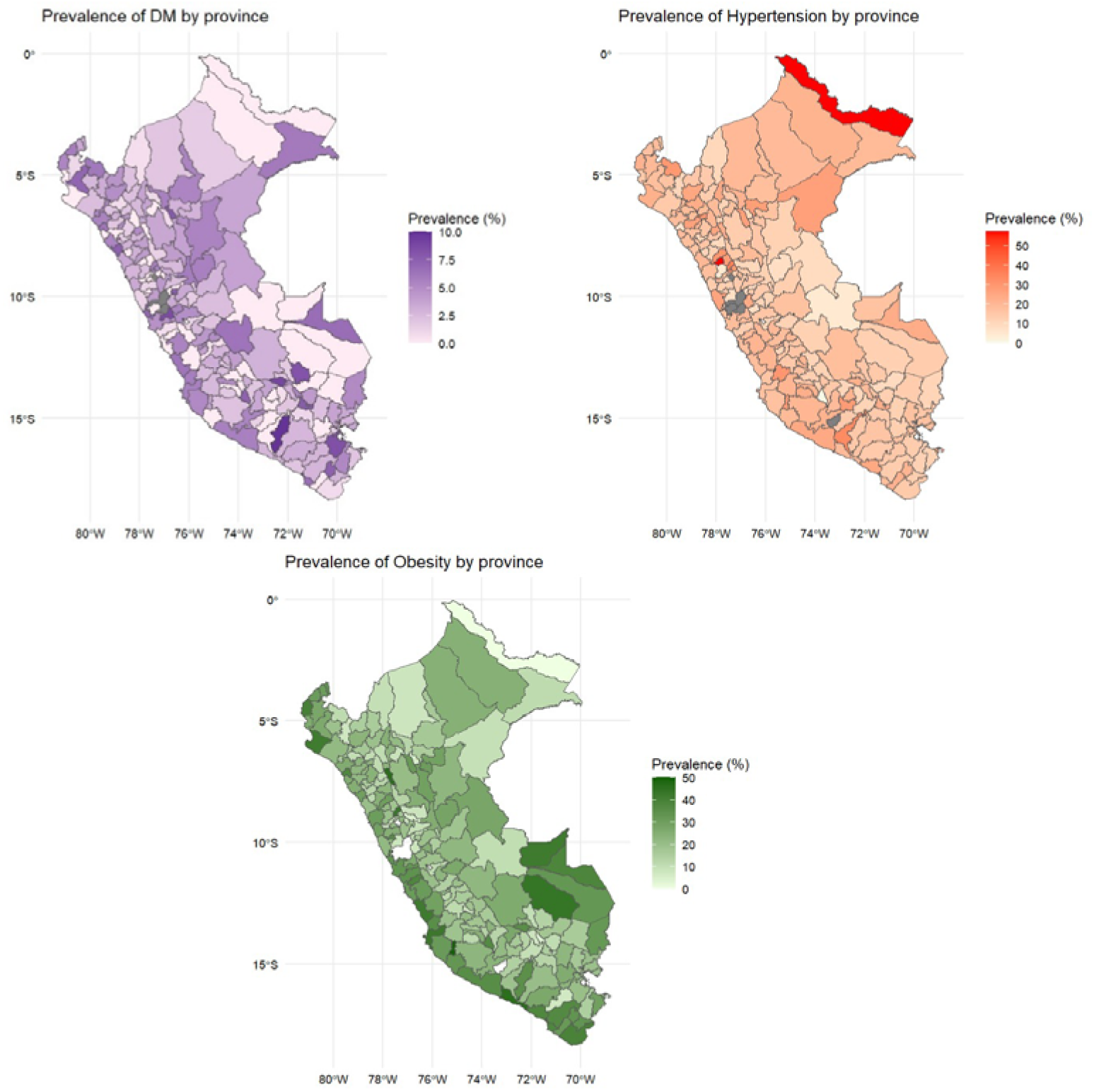
Heatmaps of the prevalences of: A) Diabetes (DM); B) Hypertension (HTN); and C) Obesity.

Figure 2 shows the Local Indicator of Spatial Association (LISA) analysis of the studied comorbidities. All results were statistically significant (p<0.001). Map A represents DM prevalence, where the provinces of Oyón (Lima) and Daniel Alcides Carrión (Pasco) show hotspots of higher diabetes prevalence, followed by Ucayali (Loreto) and Manu (Madre de Dios). In map B, we can identify that the highest prevalences of hypertension are focused in the provinces of Putumayo and Maynas (Loreto), Padre Abad (Ucayali), Pallasca, Corongo, and Sihuas (Ancash). Lastly, map C shows that hotspots of high obesity prevalence are found in Putumayo and Maynas (Loreto), Pallasca, Corongo, and Sihuas (Ancash), and Padre Abad (Ucayali).

**Figure 2.**
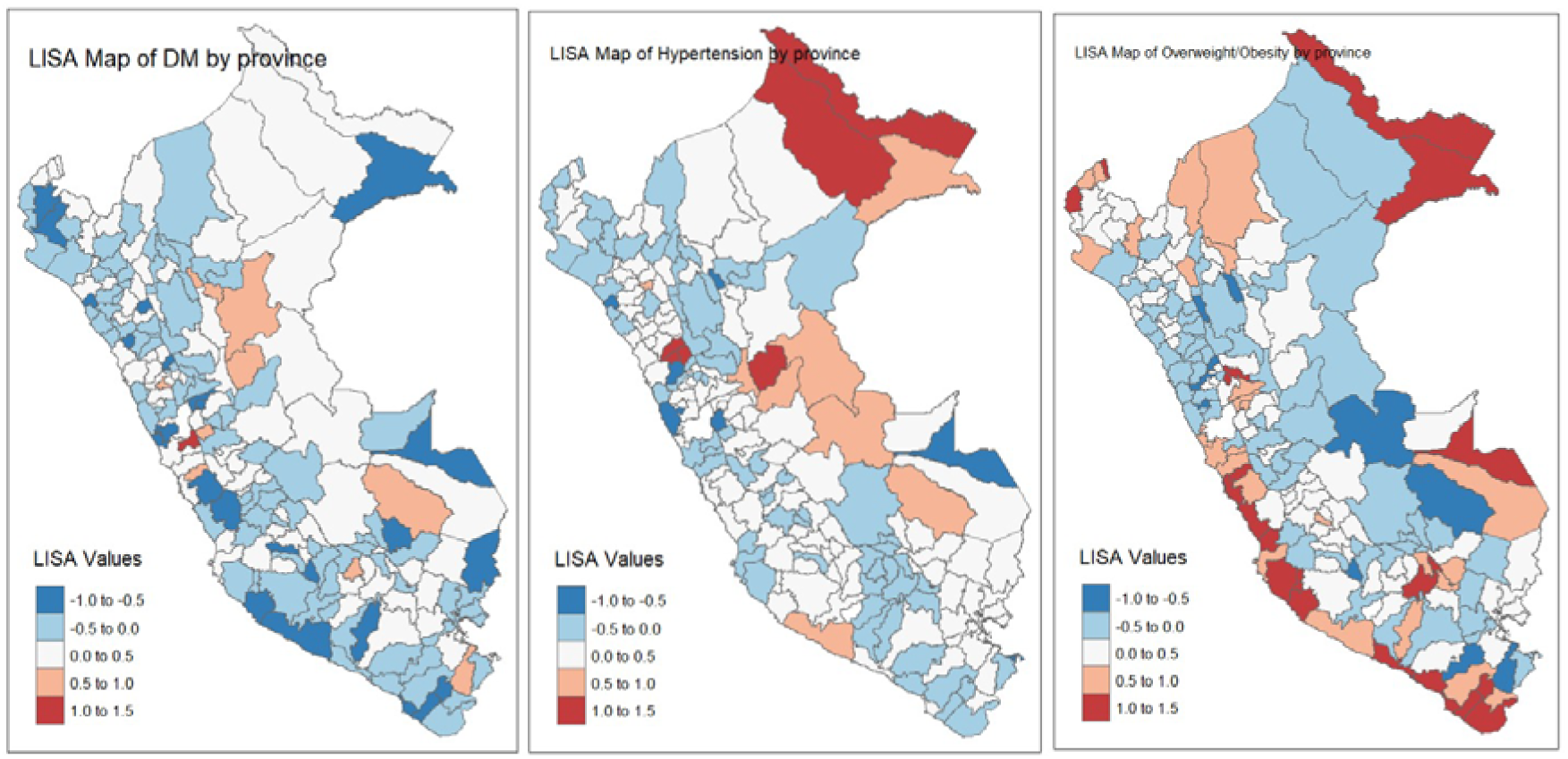
LISA maps of comorbidities: A) Diabetes (DM); B) Hypertension (HTN); and C) Obesity.

Figure 3 allows us to visualize the distribution of prevalences related to participants’ lifestyles. In Map A, with pink shades, the darkest areas represent places with the highest alcohol consumption, mainly in the provinces of Putumayo (Loreto), La Unión (Arequipa), and Sechura (Piura). Map B shows tobacco consumption prevalences in orange shades, with the highest prevalences identified in the provinces of Ucayali and Requena (Loreto), Pallasca and Corongo (Ancash), Bellavista and Mariscal Cáceres (San Martín). Finally, regarding fruit and vegetable consumption, map C shows the lightest blue shades indicating the lowest prevalences, located in the provinces of Putumayo (Loreto), Atalaya and Coronel Portillo (Ucayali), Bolognesi, Ocros, and Recuay (Ancash).

**Figure 3.**
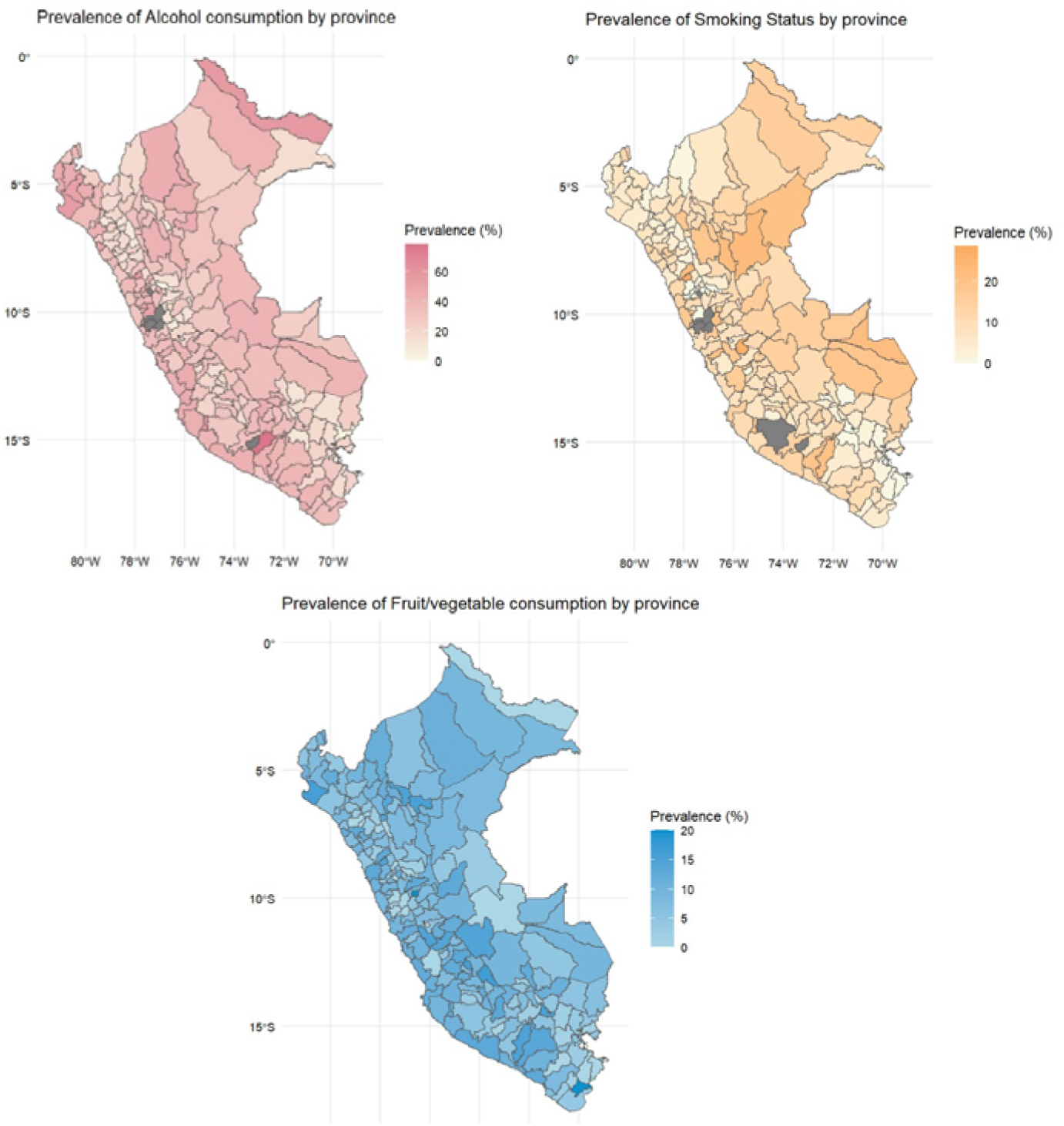
Heat maps of prevalences related to lifestyles: A) Alcohol consumption; B) Smoking status; and C) Fruit and vegetable consumption.

Figure 4 shows the LISA analysis of the dimensions of Peruvians’ lifestyles. All results were statistically significant (p<0.001). Map A shows the dispersed high prevalences of alcohol consumption, found mainly in the provinces of Talara, Paita, Piura, and Sechura (Piura), Cajamarca, Celendín, Hualgayoc, San Marcos, and San Pablo (Cajamarca); Chincha and Ica (Ica); Daniel Alcides Carrión (Pasco); Ambo and Lauricocha (Huánuco); Caraveli and La Unión (Arequipa); Putina and Azángaro (Puno). On the other hand, map B shows a focus of high prevalences of tobacco consumption in the provinces of Maynas, Requena, and Ucayali (Loreto); Bellavista, Picota, and Huallaga (San Martín); Purus and Atalaya (Ucayali); Tahuamanu and Tambopata (Madre de Dios); Melgar, Lampa, El Collao, Chuchuito, and Yunguy (Puno). Finally, map C shows a very dispersed pattern in relation to the prevalence of fruit and vegetable consumption, affecting the provinces of Huarochirí and Canta (Lima); Huamalíes (Huánuco);

**Figure 4.**
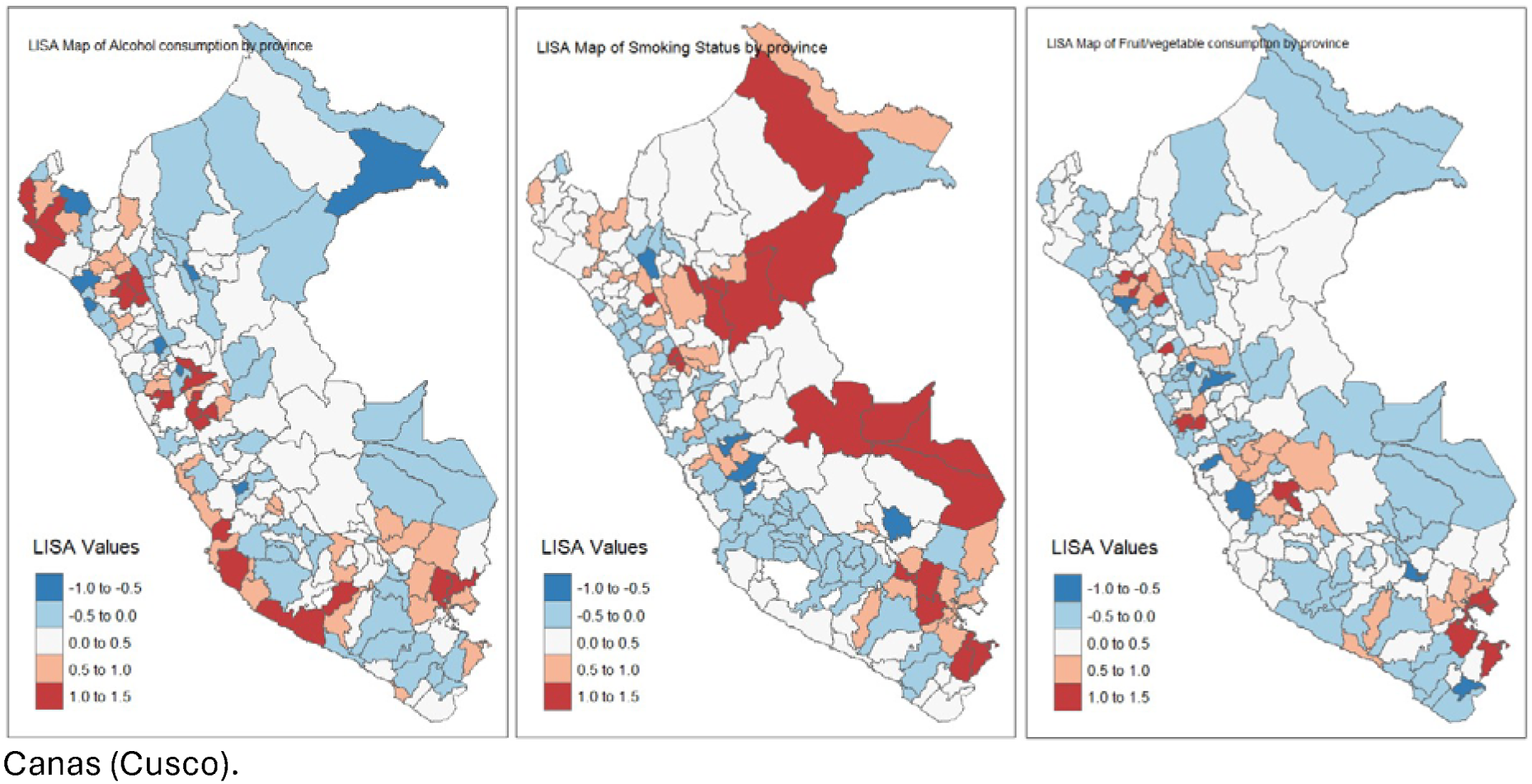
LISA spatial representation map of participants’ lifestyles: A) Alcohol consumption; B) Smoking status; and C) Fruit and vegetable consumption.

## Discussion

### Geospatial Analysis of DM, HTN, and Obesity Prevalences

Our geospatial analysis revealed distinctive patterns in the distribution of DM, HTN, and obesity at the provincial level in Peru, highlighting significant regional disparities that require attention in public health policies.

Regarding DM, the LISA analysis identified hotspots of high prevalence in diverse provinces. The provinces of Oyón (Lima) and Daniel Alcides Carrión (Pasco) showed the highest prevalences, followed by Ucayali (Loreto) and Manu (Madre de Dios). This heterogeneous distribution suggests the interaction of multiple factors in the epidemiology of DM in Peru. In urban provinces, especially in Lima, the high prevalence could be attributed to rapid urbanization and associated lifestyle changes, including Westernized diets and reduced physical activity, as noted by Bernabé-Ortiz et al. ^(16)^. The presence of high-prevalence hotspots in central highland provinces, such as Daniel Alcides Carrión, could be related to genetic, socioeconomic, and environmental factors specific to these high-altitude regions ^(17,18)^.

The high prevalence of DM in jungle provinces such as Ucayali and Manu is particularly concerning and warrants deeper analysis. These findings coincide with recent studies that have documented a significant increase in DM in Amazonian populations ^(19)^. Factors such as rapid nutritional transition, deforestation, and changes in traditional ways of life could be contributing to this phenomenon. Additionally, limited access to preventive health services and lack of early detection programs in these remote provinces could be underestimating the true magnitude of the problem ^(20)^.

Regarding HTN, our results show a concentration of high prevalence in the provinces of Putumayo and Maynas (Loreto), Padre Abad (Ucayali), Pallasca, Corongo, and Sihuas (Ancash). These findings are consistent with previous studies that have identified a higher burden of HTN in rural and jungle areas of Peru ^(4)^. The high prevalence in these provinces could be explained by a combination of dietary, environmental, and socioeconomic factors. High sodium consumption, common in these regions due to food preservation practices, could be a significant contributing factor. Additionally, psychosocial stress associated with the rapid transformation of traditional ways of life and community displacement could be influencing the prevalence of HTN ^(20)^.

Regarding obesity, our analysis revealed a more dispersed geographical distribution, with hotspots of high prevalence in provinces such as Putumayo and Maynas (Loreto), Pallasca, Corongo, and Sihuas (Ancash), and Padre Abad (Ucayali). The high prevalence in urban coastal provinces is consistent with existing literature and can be attributed to the adoption of sedentary lifestyles and high-calorie diets ^(21)^. However, the presence of high-prevalence hotspots in jungle provinces is a finding that deserves special attention. This pattern could be reflecting the rapid nutritional transition in these areas, characterized by the abandonment of traditional diets and the adoption of processed foods with high energy density ^(22,23)^.

It is important to highlight the geographical overlap observed between high prevalences of DM, HTN, and obesity, particularly in jungle provinces such as Putumayo and Maynas. This coincidence suggests the presence of common risk factors and the need for comprehensive interventions that simultaneously address these conditions. The Westernization of diet has played an important role, increasing the consumption of ultra-processed foods rich in sugars and fats, which has replaced diets rich in fiber and healthy nutrients ^(24,25)^. Climatic conditions in these provinces could also be related to limitations in performing regular physical activities, contributing to a sedentary lifestyle ^(26)^.

Our findings have important implications for public health policy in Peru. They suggest the need for differentiated and culturally adapted strategies to address the burden of chronic diseases in different provinces of the country. In particular, urgent attention is required in jungle provinces, where the convergence of high prevalence of DM, HTN, and obesity indicates an emerging health crisis.

Future research should delve deeper into the specific mechanisms that explain these geographic patterns at the provincial level, including longitudinal studies to evaluate temporal trends and multilevel analyses to examine the interaction between individual and contextual factors.

### Geospatial Analysis of Lifestyle Prevalences

Our LISA analysis of lifestyle dimensions in the Peruvian population revealed distinctive geographic patterns at the provincial level. This provided valuable information for the formulation of targeted public health policies.

Regarding alcohol consumption, a dispersed distribution of high prevalences was observed, mainly in coastal and Andean provinces. The provinces of Talara, Paita, Piura, and Sechura (Piura), Cajamarca, Celendín, Hualgayoc, San Marcos, and San Pablo (Cajamarca), Chincha and Ica (Ica), Daniel Alcides Carrión (Pasco), Ambo and Lauricocha (Huánuco), Caravelí and La Unión (Arequipa), and Putina and Azángaro (Puno) showed the highest rates of excessive alcohol consumption. These findings are consistent with previous studies that have identified higher alcohol consumption in coastal urban areas and some Andean regions of Peru ^(27)^.

The high prevalence in coastal provinces such as Talara and Paita could be associated with socioeconomic and cultural factors specific to these urban areas. Molina-Quiñones and Salazar-Taquiri ^(28)^ suggest that, in these areas, alcohol consumption is strongly linked to social and cultural events, in addition to greater availability and access to alcoholic beverages. On the other hand, the high prevalence in Andean provinces such as Celendín and Hualgayoc could be related to traditional consumption patterns and the use of alcohol in rituals and community celebrations ^(29)^.

Regarding tobacco consumption, our analysis revealed a concentration of high prevalences in jungle provinces and some Andean areas. The provinces of Maynas, Requena, and Ucayali (Loreto), Bellavista, Picota, and Huallaga (San Martín), Purús and Atalaya (Ucayali), Tahuamanu and Tambopata (Madre de Dios), and Melgar, Lampa, El Collao, Chucuito, and Yunguyo (Puno) presented the highest smoking rates. These results reflect a complex pattern that differs from the typical distribution of tobacco consumption in the country.

The high prevalence in jungle provinces such as Maynas and Requena could be explained by cultural and economic factors specific to these regions. The reasons behind this may be due to the fact that, in these areas, tobacco is still used in traditional rituals and medicinal practices. Additionally, the lower penetration of anti-smoking campaigns in these remote regions could contribute to the persistence of high consumption rates ^(29)^.

Regarding fruit and vegetable consumption, our analysis showed a dispersed pattern of low consumption, affecting coastal, highland, and jungle provinces. The provinces of Huarochirí and Canta (Lima), Huamalíes (Huánuco), and Canas (Cusco) presented the lowest prevalences of adequate fruit and vegetable consumption. This heterogeneous distribution suggests the influence of multiple factors on the dietary patterns of the Peruvian population.

The low intake of fruits and vegetables in provinces such as Huarochirí and Canta, close to Lima, might seem paradoxical given their proximity to a large urban center. However, this phenomenon could be explained by economic and accessibility factors. It has been noted that, despite the proximity to wholesale markets, the high cost of fruits and vegetables relative to local incomes may limit their consumption in these peri-urban areas ^(30)^.

### Limitations of the Study

The main limitations that should be considered in this study for a correct interpretation of results are as follows. First, secondary databases may generate biases related to the lack of specific and detailed information on study variables that were not recorded in the original database. Second, for the DM variable, only self-report has been considered, given that there is no laboratory analysis in the DHS.

## Conclusions

This study reveals complex geographical patterns and significant disparities in the prevalence of diabetes mellitus, hypertension, obesity, and unhealthy lifestyles among Peruvian provinces. Hotspots of high prevalence of these chronic conditions were identified mainly in jungle regions and some coastal and Andean areas, with notable overlap in several Amazonian provinces. Lifestyle patterns, including alcohol consumption, tobacco use, and fruit and vegetable intake, showed heterogeneous distributions that reflect the cultural and socioeconomic diversity of the country. The influence of Westernization on dietary and life habits was evident, particularly in urban and transitional areas. These findings underscore the complex interaction between geographical, socioeconomic, and cultural factors in determining the health profiles of the Peruvian population, highlighting the vulnerability of certain regions, especially jungle provinces and some Andean ones, where high prevalences of chronic diseases and unhealthy lifestyles converge.

Based on these findings, it is recommended to develop and implement differentiated public health policies that are culturally adapted to the specific realities of each province. It is crucial to strengthen primary care systems in areas identified with a high burden of chronic diseases, prioritizing early detection and proper management. Culturally appropriate healthy lifestyle promotion programs should be implemented, simultaneously addressing the social determinants of health through intersectoral policies. It is essential to promote additional research to better understand the factors contributing to the observed geographic disparities, establish robust surveillance systems at the provincial level, and foster intersectoral collaboration and community empowerment in the design and implementation of health interventions. Finally, priority should be given to reducing health inequalities, directing specific resources and programs to the most vulnerable provinces and populations identified in this study, with the aim of improving health equity throughout the country.

### Authors’ contribution

Fiorella E. Zuzunaga-Montoya: Conceptualization, Investigation, Methodology, Resources, Writing - Original Draft, Writing - Review & Editing

Luisa Erika Milagros Vásquez-Romero: Investigation, Project administration, Writing - Original Draft, Writing - Review & Editing

Joan A. Loayza-Castro: Investigation, Resources, Writing - Original Draft, Writing - Review & Editing

Jhonatan Roberto Astucuri Hidalgo: Software, Data Curation, Formal analysis, Writing - Review & Editing

Lupita Ana Maria Valladolid-Sandoval: Validation, Visualization, Writing - Original Draft, Writing - Review & Editing

Víctor Juan Vera-Ponce: Methodology, Supervision, Funding acquisition, Writing - Review & Editing

## Acknowledgments

A special thanks to the members of Universidad Nacional Toribio Rodríguez de Mendoza de Amazonas (UNTRM), Amazonas, Peru for their support and contributions throughout the completion of this research.

## Informed consent

The primary study from which the database was obtained provided the required informed consent, however, for the present study it was not required.

## Data availability

The data supporting the findings of this study can be accessed by the original research paper at the follow link: https://proyectos.inei.gob.pe/microdatos/

## Bibliographic references

1. World Health Organization. Noncommunicable diseases: what ministries of finance, tax and revenue need to know [Internet]. Geneva: World Health Organization; 2016 [citado el 26 de noviembre de 2021]. Disponible en: https://apps.who.int/iris/handle/10665/250227

2. Vera-Ponce VJ, Zuzunaga-Montoya FE, Romero LEMV, Castro JAL, Carrillo CIG de, Chenet SM. Lifestyle patterns in the Peruvian population: analysis of tobacco, alcohol and fruit/vegetable consumption based on a nine-year national survey. [Internet]. medRxiv; 2024 [citado el 27 de mayo de 2024]. p. 2024.05.26.24307960. doi:10.1101/2024.05.26.24307960

3. Carrillo-Larco R, Bernabé-Ortiz A. Diabetes mellitus tipo 2 en Perú: una revisión sistemática sobre la prevalencia e incidencia en población general. Rev Peru Med Exp Salud Publica. 2019;36(1):26–36. doi:10.17843/rpmesp.2019.361.4027

4. Ruiz-Alejos A, Carrillo-Larco RM, Bernabé-Ortiz A. Prevalence and incidence of arterial hypertension in Peru: a systematic review and meta-analysis. Rev Peru Med Exp Salud Publica. 2021;38(4):521–9. doi:10.17843/rpmesp.2021.384.8502

5. Adams KJ, Chirinos JL. Prevalencia de factores de riesgo para síndrome metabólico y sus componentes en usuarios de comedores populares en un distrito de Lima, Perú. Revista Peruana de Medicina Experimental y Salud Pública. 2018;35(1):39–45. doi:10.17843/rpmesp.2018.351.3598

6. Miranda JJ, Gilman RH, Smeeth L. Differences in cardiovascular risk factors in rural, urban and rural-to-urban migrants in Peru. Heart. 2011;97(10):787–96. doi:10.1136/hrt.2010.218537

7. Centro Nacional de Alimentación y Nutrición. Estado Nutricional En Adultos de 18 a 59 Años VIANEV 2017–2018. Instituto Nacional de Salud: Lima, Peru. 2021;191.

8. Soto A. Barreras para una atención eficaz en los hospitales de referencia del Ministerio de Salud del Perú: atendiendo pacientes en el siglo XXI con recursos del siglo XX. Revista Peruana de Medicina Experimental y Salud Publica. 2019;36(2):304–11. doi:10.17843/rpmesp.2019.362.4425

9. Del Castillo-Fernández D, Brañez-Condorena A, Villacorta-Landeo P, Saavedra-García L, Bernabé-Ortiz A, Miranda J, et al. Avances en la investigación de enfermedades crónicas no transmisibles en el Perú. Anales de la Facultad de Medicina. 2020;81(4):444–52. doi:10.15381/anales.v81i4.18798

10. Perú: Encuesta Demográfica y de Salud Familiar, DHS 2023 [Internet]. [citado el 5 de junio de 2024]. Disponible en: https://www.gob.pe/institucion/inei/informes-publicaciones/5601739-peru-encuesta-demografica-y-de-salud-familiar-DHS-2023

11. NCD Risk Factor Collaboration (NCD-RisC). Worldwide trends in blood pressure from 1975 to 2015: a pooled analysis of 1479 population-based measurement studies with 19·1 million participants. Lancet. 2017;389(10064):37–55. doi:10.1016/S0140-6736(16)31919-5

12. Wechsler H, Isaac N. “Binge” Drinkers at Massachusetts Colleges: Prevalence, Drinking Style, Time Trends, and Associated Problems. JAMA. 1992;267(21):2929–31. doi:10.1001/jama.1992.03480210091038

13. Wechsler H, Nelson TF. Binge drinking and the American college student: what’s five drinks? Psychol Addict Behav. 2001;15(4):287–91. doi:10.1037//0893-164x.15.4.287

14. Wang DD, Li Y, Bhupathiraju SN, Rosner BA, Sun Q, Giovannucci EL, et al. Fruit and Vegetable Intake and Mortality. Circulation. 2021;143(17):1642–54. doi:10.1161/CIRCULATIONAHA.120.048996

15. Obesity and overweight [Internet]. [citado el 27 de julio de 2024]. Disponible en: https://www.who.int/news-room/fact-sheets/detail/obesity-and-overweight

16. Bernabé-Ortiz A, Carrillo-Larco RM, Gilman RH, Miele CH, Checkley W, Wells JC, et al. Geographical variation in the progression of type 2 diabetes in Peru: The CRONICAS Cohort Study. Diabetes Res Clin Pract. 2016;121:135–45. doi:10.1016/j.diabres.2016.09.007

17. Asenjo-Alarcón JA, Oblitas-Gonzales A, Asenjo-Alarcón JA, Oblitas-Gonzales A. Complicaciones crónicas microvasculares en usuarios con diabetes mellitus tipo 2 de una ciudad andina del Perú. Revista de Salud Pública. 2022;24(3):1. doi:10.15446/rsap.v24n3.100418

18. López Gregorio E, Belanche Bartolomé J, Elizondo Lugo M. Revisión sistemática sobre el efecto de la modificación del estilo de vida en diabetes mellitus de tipo 2 en pacientes ancianos. Revista Sanitaria de Investigación. 2021;2(11 (Noviembre)):96.

19. Rocca J, Calderón M, La Rosa A, Seclén S, Castillo O, Pajuelo J, et al. Type 2 diabetes mellitus in Peru: A literature review including studies at high-altitude settings. Diabetes Res Clin Pract. 2021;182:109132. doi:10.1016/j.diabres.2021.109132

20. Bernabé-Ortiz A, Carrillo-Larco RM, Gilman RH, Checkley W, Smeeth L, Miranda JJ, et al. Contribution of modifiable risk factors for hypertension and type-2 diabetes in Peruvian resource-limited settings. J Epidemiol Community Health. 2016;70(1):49–55. doi:10.1136/jech-2015-205988

21. Pajuelo Ramírez J, Torres Aparcana L, Agüero Zamora R, Pajuelo Ramírez J, Torres Aparcana L, Agüero Zamora R. Asociación entre obesidad abdominal y factores demográficos, según niveles de altitud en el Perú. Anales de la Facultad de Medicina. 2020;81(2):167–73. doi:10.15381/anales.v81i2.18408

22. Tejada López YO, Choquehuanca Zambrano GM, Goicochea Ríos E del S, Vicuña Villacorta JE, Guzmán Aybar OY, Tejada López YO, et al. Perfil clínico-epidemiológico del síndrome metabólico en adultos atendidos en el hospital I Florencia de Mora EsSALUD. Horizonte Médico (Lima) [Internet]. 2020 [citado el 10 de agosto de 2024];20(4). doi:10.24265/horizmed.2020.v20n4.07

23. Contreras-Pulache H, Pérez-Campos P, Huapaya-Huertas O, Chacón-Torrico H, Champin- Mimbela D, Freyre-Adrianzen L, Arévalo-León C, Torres-Llaque S, Black-Tam C. La salud en las comunidades nativas amazónicas del Perú. Rev Peru Epidemiol. 2014 Abr;18(1):91-102.

24. Clemente-Suárez VJ, Beltrán-Velasco AI, Redondo-Flórez L, Martín-Rodríguez A, Tornero-Aguilera JF. Global Impacts of Western Diet and Its Effects on Metabolism and Health: A Narrative Review. Nutrients. 2023;15(12):2749. doi:10.3390/nu15122749

25. Pineda E, Stockton J, Scholes S, Lassale C, Mindell JS. Food environment and obesity: a systematic review and meta-analysis. BMJ Nutrition, Prevention & Health [Internet]. 2024 [citado el 10 de agosto de 2024];7(1). doi:10.1136/bmjnph-2023-000663

26. Tompkins AM, Caporaso L, Biondi R, Bell JP. A Generalized Deforestation and Land-Use Change Scenario Generator for Use in Climate Modelling Studies. PLoS One. 2015;10(9):e0136154. doi:10.1371/journal.pone.0136154

27. Gonález Vázquez A, López García CS, Tizoc Marquez A. Modelo explicativo sobre la conducta de consumo de alcohol de jóvenes del área rural y urbana. Ra Ximhai: revista científica de sociedad, cultura y desarrollo sostenible. 2020;16(Extra 3):235–50.

28. Molina-Quiñones H, Salazar-Taquiri V, Molina-Quiñones H, Salazar-Taquiri V. Factores asociados al consumo de alcohol en adolescentes residentes en Lima, Perú. Revista Habanera de Ciencias Médicas [Internet]. 2022 [citado el 10 de agosto de 2024];21(3). Disponible en: http://scielo.sld.cu/scielo.php?script=sci_abstract&pid=S1729-519X2022000300011&lng=es&nrm=iso&tlng=es

29. Consumo de alcohol, tabaco y marihuana en adolescentes de secundaria de la zona rural. Ciencia Latina. 2022;6(1):835–48. doi:10.37811/cl_rcm.v6i1.1798

30. Martínez-Velarde D, Málaga-Chávez R, Bernabe-Ortiz A. Consumo de bebidas azucaradas, verduras y frutas en sujetos con alteración del metabolismo de la glucosa. Revista Española de Nutrición Humana y Dietética. 2021;25(3):326–36. doi:10.14306/renhyd.25.3.1258

